# Baseline characteristics, management, and outcomes of 55,270 children and adolescents diagnosed with COVID-19 and 1,952,693 with influenza in France, Germany, Spain, South Korea and the United States: an international network cohort study

**DOI:** 10.1101/2020.10.29.20222083

**Authors:** Talita Duarte-Salles, David Vizcaya, Andrea Pistillo, Paula Casajust, Anthony G. Sena, Lana Yin Hui Lai, Albert Prats-Uribe, Waheed-Ul-Rahman Ahmed, Thamir M Alshammari, Heba Alghoul, Osaid Alser, Edward Burn, Seng Chan You, Carlos Areia, Clair Blacketer, Scott DuVall, Thomas Falconer, Sergio Fernandez-Bertolin, Stephen Fortin, Asieh Golozar, Mengchun Gong, Eng Hooi Tan, Vojtech Huser, Pablo Iveli, Daniel R. Morales, Fredrik Nyberg, Jose D. Posada, Martina Recalde, Elena Roel, Lisa M. Schilling, Nigam H. Shah, Karishma Shah, Marc A. Suchard, Lin Zhang, Ying Zhang, Andrew E. Williams, Christian G. Reich, George Hripcsak, Peter Rijnbeek, Patrick Ryan, Kristin Kostka, Daniel Prieto-Alhambra

## Abstract

**Objectives:** To characterize the demographics, comorbidities, symptoms, in-hospital treatments, and health outcomes among children/adolescents diagnosed or hospitalized with COVID-19. Secondly, to describe health outcomes amongst children/adolescents diagnosed with previous seasonal influenza.

**Design:** International network cohort.

**Setting:** Real-world data from European primary care records (France/Germany/Spain), South Korean claims and US claims and hospital databases.

**Participants:** Diagnosed and/or hospitalized children/adolescents with COVID-19 at age <18 between January and June 2020; diagnosed with influenza in 2017-2018.

**Main outcome measures:** Baseline demographics and comorbidities, symptoms, 30-day in-hospital treatments and outcomes including hospitalization, pneumonia, acute respiratory distress syndrome (ARDS), multi-system inflammatory syndrome (MIS-C), and death.

**Results:** A total of 55,270 children/adolescents diagnosed and 3,693 hospitalized with COVID-19 and 1,952,693 diagnosed with influenza were studied.

Comorbidities including neurodevelopmental disorders, heart disease, and cancer were all more common among those hospitalized vs diagnosed with COVID-19. The most common COVID-19 symptom was fever. Dyspnea, bronchiolitis, anosmia and gastrointestinal symptoms were more common in COVID-19 than influenza.

In-hospital treatments for COVID-19 included repurposed medications (<10%), and adjunctive therapies: systemic corticosteroids (6.8% to 37.6%), famotidine (9.0% to 28.1%), and antithrombotics such as aspirin (2.0% to 21.4%), heparin (2.2% to 18.1%), and enoxaparin (2.8% to 14.8%).

Hospitalization was observed in 0.3% to 1.3% of the COVID-19 diagnosed cohort, with undetectable (N<5 per database) 30-day fatality. Thirty-day outcomes including pneumonia, ARDS, and MIS-C were more frequent in COVID-19 than influenza.

**Conclusions:** Despite negligible fatality, complications including pneumonia, ARDS and MIS-C were more frequent in children/adolescents with COVID-19 than with influenza. Dyspnea, anosmia and gastrointestinal symptoms could help differential diagnosis. A wide range of medications were used for the inpatient management of pediatric COVID-19.

**What is already known on this topic?:**

- Most of the early COVID-19 studies were targeted at adult patients, and data concerning children and adolescents are limited.
- Clinical manifestations of COVID-19 are generally milder in the pediatric population compared with adults.
- Hospitalization for COVID-19 affects mostly infants, toddlers, and children with pre-existing comorbidities.

**What this study adds:**

⍰ This study comprehensively characterizes a large international cohort of pediatric COVID-19 patients, and almost 2 million with previous seasonal influenza across 5 countries.
⍰ Although uncommon, pneumonia, acute respiratory distress syndrome (ARDS) and multi-system inflammatory syndrome (MIS-C) were more frequent in children and adolescents diagnosed with COVID-19 than in those with seasonal influenza.
⍰ Dyspnea, bronchiolitis, anosmia and gastrointestinal symptoms were more frequent in COVID-19, and could help to differentiate pediatric COVID-19 from influenza.
⍰ A plethora of medications were used during the management of COVID-19 in children and adolescents, with great heterogeneity in the use of antiviral therapies as well as of adjunctive therapies.

## INTRODUCTION

Since January 2020, a growing number of infections by the severe acute respiratory syndrome coronavirus 2 (SARS-CoV-2) has led to an unprecedented pressure on healthcare systems worldwide. As of 14^th^ of October 2020, there were 38,002,699 confirmed cases of coronavirus disease 2019 (COVID-19) and 1,083,234 deaths according to the World Health Organization (WHO) Dashboard. COVID-19 affects all age groups, including the pediatric population, which represents 3.7% of reported cases [1].

With day-care centers and schools re-opening in many parts of the world despite high levels of community transmission, it is of considerable importance to elucidate the baseline clinical features and health outcomes seen in children/adolescents (aged below 18 years) diagnosed with COVID-19 during the first wave of the pandemic. The pediatric presentation of COVID-19 reported to date ranges from completely asymptomatic but test positive to symptoms of acute upper respiratory tract infection and rarely various severe manifestations.[2] Clinical manifestations of COVID-19 are generally milder in the pediatric population,[3] with better outcomes and lower mortality rates than adults.[4] Nevertheless, there is evidence of children/adolescents with COVID-19 requiring hospitalization and intensive care unit (ICU)-level care. Reports from the United States (US) revealed that a low number of pediatric COVID-19 cases were hospitalized (5.7%), mainly among adolescents aged 15-17 years (32%) and those with underlying comorbidities such as chronic lung disease (including asthma), cardiovascular disease, or immunosuppression.[5] These findings are in line with a report on pediatric COVID-19 patients (<16 years old) in China, where only 1.8% were admitted to the ICU.[6] A study conducted in 25 European countries, on the other hand, found that in a sample of 582 children/adolescents aged below 18 years, 63% were hospitalized.[7]

To date, most clinical guidelines recommend supportive care as the mainstay of therapy in children [8-10], but there is little data to recommend or reject the use of specific immunomodulatory drugs or antivirals. It also remains to be elucidated whether children/adolescents show a different clinical presentation.[11] We conducted a literature review for articles published in PubMed and Medrxiv (preprints) between December 2019 and June 2020 that reported on patients with a confirmed COVID-19 diagnosis. Of the 1320 studies that met the inclusion criteria, only 79 studies were on children/adolescents, most of which (63%) were local case reports or case series. This compellingly demonstrates a large remaining gap in existing efforts to define the characteristics of the pediatric population in a real-world setting at a large scale.

In this study, we aimed to describe the demographics, comorbidities, symptoms, in-hospital treatments, and health outcomes of children/adolescents diagnosed or hospitalized with COVID-19, using electronic health records (EHRs) and health claims databases across the US, Europe and Asia. In addition, we compared these cohorts with children/adolescents diagnosed with seasonal influenza in 2017-2018 as a benchmark.

## METHODS

### Study design, setting and data sources

This study is part of the Characterizing Health Associated Risks, and Your Baseline Disease In SARS-COV-2 (CHARYBDIS) study, a large-scale multinational cohort study using routinely-collected primary care and hospital EHRs, hospital billing data and insurance claims data from the US, Europe (the Netherlands, Spain, the UK, Germany and France) and Asia (South Korea and China).

From the nineteen databases contributing data to CHARYBDIS, only those with data on patients below the age of 18 years with a clinical diagnosis of COVID-19 or a SARS-CoV-2 positive test between January and June 2020 were included. A cohort of children/adolescents diagnosed with seasonal influenza in 2017-2018 was included for comparison.

To be included in the study, databases had to have a minimum of 140 children/adolescents (aged below 18 years). This cut-off was deemed necessary to estimate with sufficient precision (confidence interval width of ±5%) the prevalence of a previous condition or 30-day risk of an outcome affecting 10% of the study population. Data results for this paper were extracted from CHARYBDIS results on the 22^nd^ of September 2020. Figure 1 represents the selection process of the databases for this study. Eleven databases fulfilled the inclusion criteria: STARR-OMOP (US)[12], CU-AMC HDC (US), HealthVerity (US), CUIMC (US), OPTUM-EHR (US), Premier (US), IQVIA-OpenClaims (US), IQVIA-LPD France (France), IQVIA-DA Germany (Germany), SIDIAP (Spain)[13], HIRA (South Korea). Among these, five databases contributed to the hospitalized cohort: Premier, OPTUM-EHR, IQVIA-OpenClaims, CUIMC, HIRA. A more detailed description of the included data sources is available in Supplementary Table 1.

**Figure 1.**
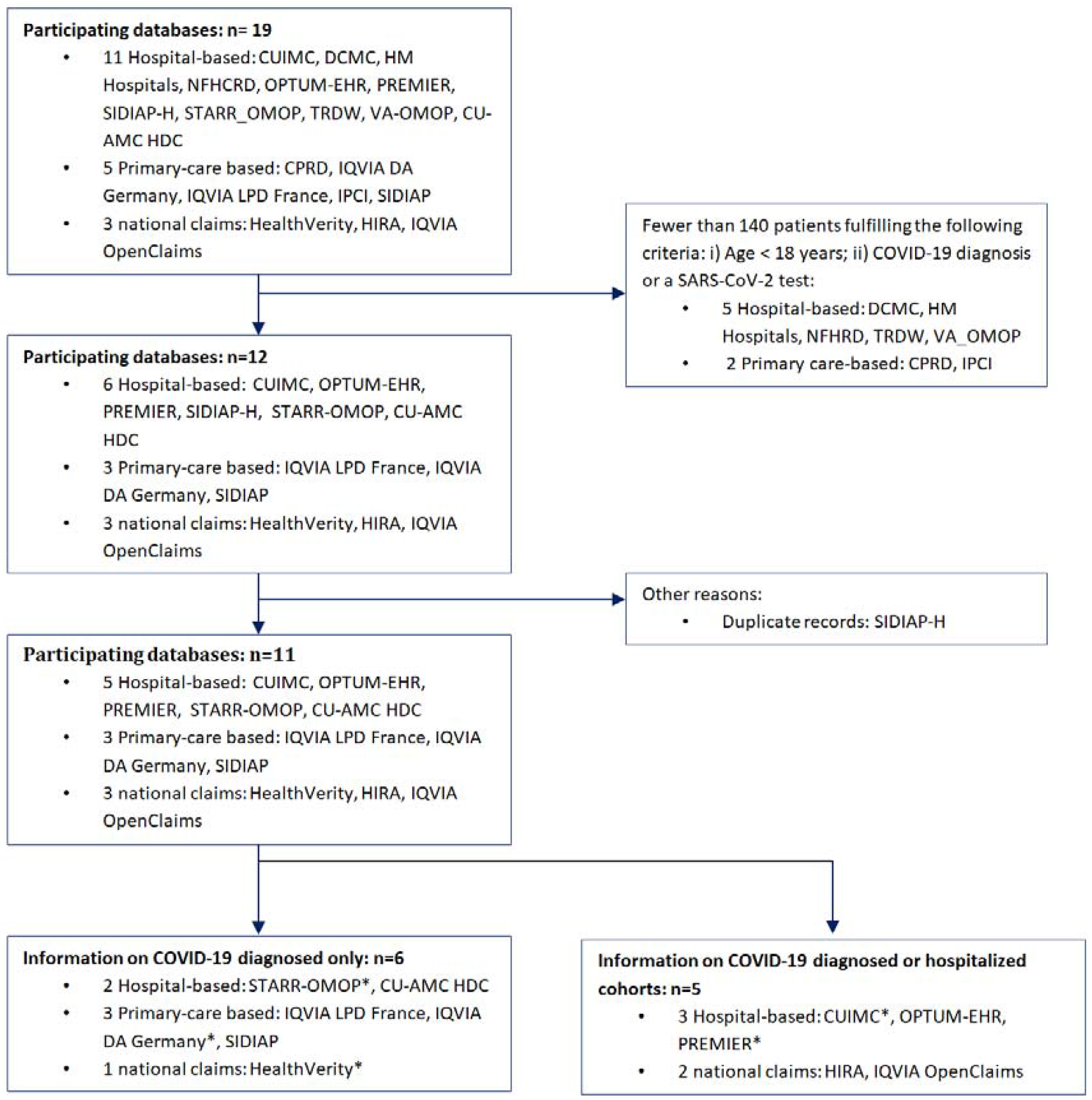
Database selection process. Colorado University Anschuz Medical Campus Health Data Compass (CU-AMC HDC), Columbia University Irving Medical Center (CUIMC), Clinical Practice Research Datalink (CPRD), Data Analyzer (DA), Daegu Catholic University Medical Center (DCMC), Health Insurance Review & Assessment Service (HIRA), Integrated Primary Care Information (IPCI), Longitudinal Patient Data (LPD), Nanfang Hospital COVID-19 Research Database (NFHCRD), Information System for Research in Primary Care (SIDIAP), STAnford medicine Research data Repository (STARR-OMOP), Tufts Research Data Warehouse (TRDW), Department of Veterans Affairs (VA-OMOP) *No prior observation time available and therefore excluded from description of comorbidities.

### Study participants and follow-up

Two non-mutually exclusive cohorts were included: 1) children/adolescents with COVID-19 diagnosis or a SARS-CoV-2 positive test (index date was the first of the two events), 2) children/adolescents hospitalized (index date was hospitalization date), with COVID-19 diagnosis or a SARS-CoV-2 positive test 21 days before or after hospitalization date. A similar diagnosed cohort of children/adolescents with seasonal influenza diagnosis or positive influenza test in 2017-2018 was also included. Study participants could contribute information to all cohorts (diagnosed or hospitalized with COVID-19, or diagnosed with influenza). Individuals diagnosed with COVID-19 could be part of the hospitalized cohort if COVID-19 was diagnosed during or at the time or 21 days before hospitalization. Cohort participants were followed in each cohort from the index date to the earliest of death, end of the observation period, or 30 days after. The codes used to identify COVID-19 are described in Supplementary Table 2. In order to describe history of comorbidities, only participants with at least 365 days of prior observation before index date were included. Children below age one were excluded from the cohorts requiring 365 days of prior observation.

### Baseline characteristics, symptoms, drug use and outcomes of interest

Baseline information on age at index date and conditions up to 1 year before index date were identified. Conditions were ascertained based on the Systematized Nomenclature of Medicine – Clinical Terms (SNOMED CT) hierarchy, with all descendant codes included. Detailed definitions of each condition can be consulted in Supplementary Table 2.

Symptoms recorded at index date included fever, cough, dyspnea, malaise or fatigue, myalgia, anosmia or hyposmia or dysgeusia, gastrointestinal tract symptoms, diarrhea, vomits and nausea. All drugs prescribed/dispensed during the 30-days follow-up after the index date were ascertained. Individual medications were categorized using Anatomical Therapeutic Chemical (ATC) groupings. For the study of medications potentially used for COVID-19, we assessed all medications included in at least two randomized controlled trials [14]. The list was further enriched with suggestions from key stakeholders including regulatory agencies, key opinion leaders, and pharma industry. Medicines of interest were grouped into: 1) repurposed medications- those with alternative indications but believed to be efficacious as antivirals, 2) adjuvant therapies - used allegedly for the treatment or prevention of COVID-19 complications [15]. All conditions and medications and additional time windows (a month prior and on index date) are reported in full and are available in an interactive website: https://data.ohdsi.org/Covid19CharacterizationCharybdis/.

The 30-day outcomes described in the diagnosed cohorts included hospitalization, death, pneumonia, and multi-system inflammatory syndrome in children (MIS-C, Kawasaki disease, or toxic shock syndrome). In the hospitalized cohorts, we additionally report sepsis, total cardiovascular disease events, acute respiratory disease syndrome (ARDS), cardiac arrhythmia, and bleeding. The definition of each outcome is provided in Supplementary Table 2.

### Statistical analyses

All data were standardized to the Observational Medical Outcomes Partnership (OMOP) Common Data Model (CDM).[16] A common analytical code for the CHARYBDIS study was developed for the OHDSI Methods library which was run locally in each database. Only aggregate results from each database were publicly shared. The CHARYBDIS protocol and source code can be found at https://github.com/ohdsi-studies/Covid19CharacterizationCharybdis. Results were extracted from CHARYBDIS on October 1st 2020.

Demographics, history of comorbidities, symptoms and outcomes were summarized as proportions, calculated by dividing the number of people within a given category by the total number of people. The proportion of users of each medication was determined for the hospitalized children/adolescents as the percentage of subjects who had >=1 day during the time window overlapping with a drug use period for each medication or drug class of interest. Utilization of repurposed and adjuvant drugs up to 30 days after admission was described.

The distribution of conditions a year prior to index, symptoms at index, outcomes and medications up to 30 days post-index date in COVID-19 diagnosed cohort were compared to the hospitalized COVID-19 or the diagnosed influenza cohorts. Standardized mean differences (SMD) were calculated when comparing the characteristics of study cohorts.

We performed a sensitivity analysis describing characteristics of cases with no prior observation time in order to understand the impact of lack of prior observation time in the results.

We used R version 3.6 (R Foundation for Statistical Computing, Vienna, Austria) for data visualization. All the data partners obtained Institutional Review Board (IRB) approval or exemption to conduct this study, as required.

## RESULTS

A total of 55,270 children/adolescents diagnosed (3,693 hospitalized) with COVID-19, and 1,956,358 diagnosed with seasonal influenza were included. Data were obtained from 5 US hospital EHR databases (STARR-US [with the pediatric population representing 3.9% of all COVID-19 cases in this database], CU-AMC-HDC-US [2.6% of all COVID-19 cases], CUIMC-US [4.1% of all COVID-19 cases], OPTUM-EHR-US [4.4% of all COVID-19 cases] and PREMIER-US [3.9% of all COVID-19 cases]), 3 European primary care records databases (IQVIA-LPD-France [4.0% of all COVID-19 cases], IQVIA-DA-Germany [7.1% of all COVID-19 cases], SIDIAP-Spain [3.7% of all COVID-19 cases]), and 2 US (HealthVerity-US [4.6% of all COVID-19 cases], IQVIA-OpenClaims-US [2.9% of all COVID-19 cases]) and 1 Asian claims databases (HIRA South Korea [3.3% of all COVID-19 cases]). Up to at least 1 year of pre-index observation time was available only in CU-AMC-HDC-US, OPTUM-EHR-US, SIDIAP-Spain, and IQVIA-OpenClaims-US. Figure 1 shows a flowchart outlining the reasons for exclusion of additional 8 data sources available in CHARYBDIS.

### Demographics

Age at diagnosis of COVID-19 varied across regions. In SIDIAP-Spain, CUIMC-US and STARR-US the majority of children with COVID-19 were diagnosed at ages 0 to 4 years (around one third), while the proportion of children from 0 to 4 years was only 11.4% and 11.6% in IQVIA-LPD-France and CUIMC-US, respectively. More consistently, most of the hospital admissions were seen in the younger groups (0-4 years), e.g. 40.6% of those hospitalized in IQVIA-OpenClaims-US, and 76.2% in Premier-US. Male gender was more common in all databases except for IQVIA-LPD-France (46.8% male) and STARR-US (49.2%) (Table 1).

**Table 1.**
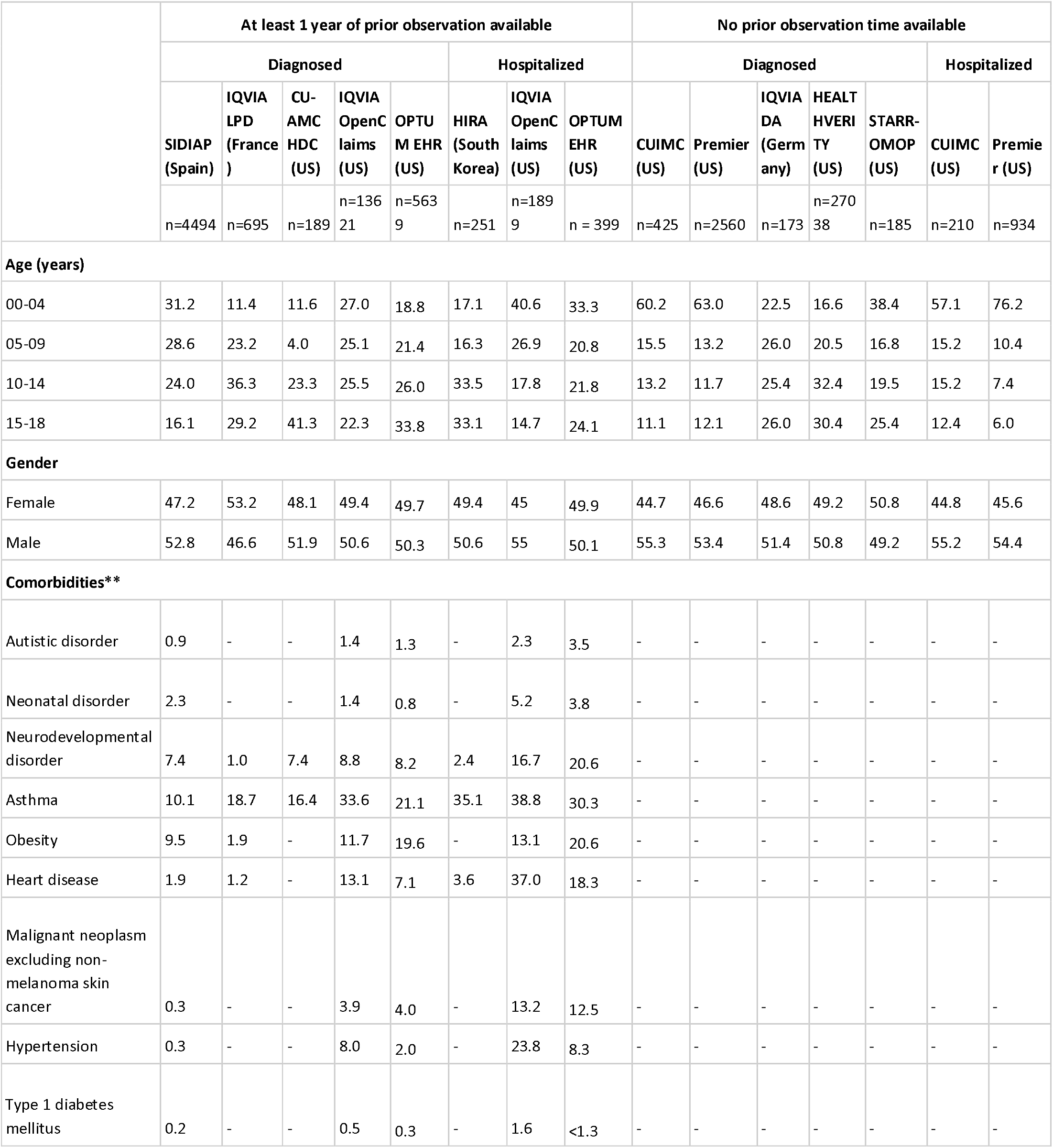
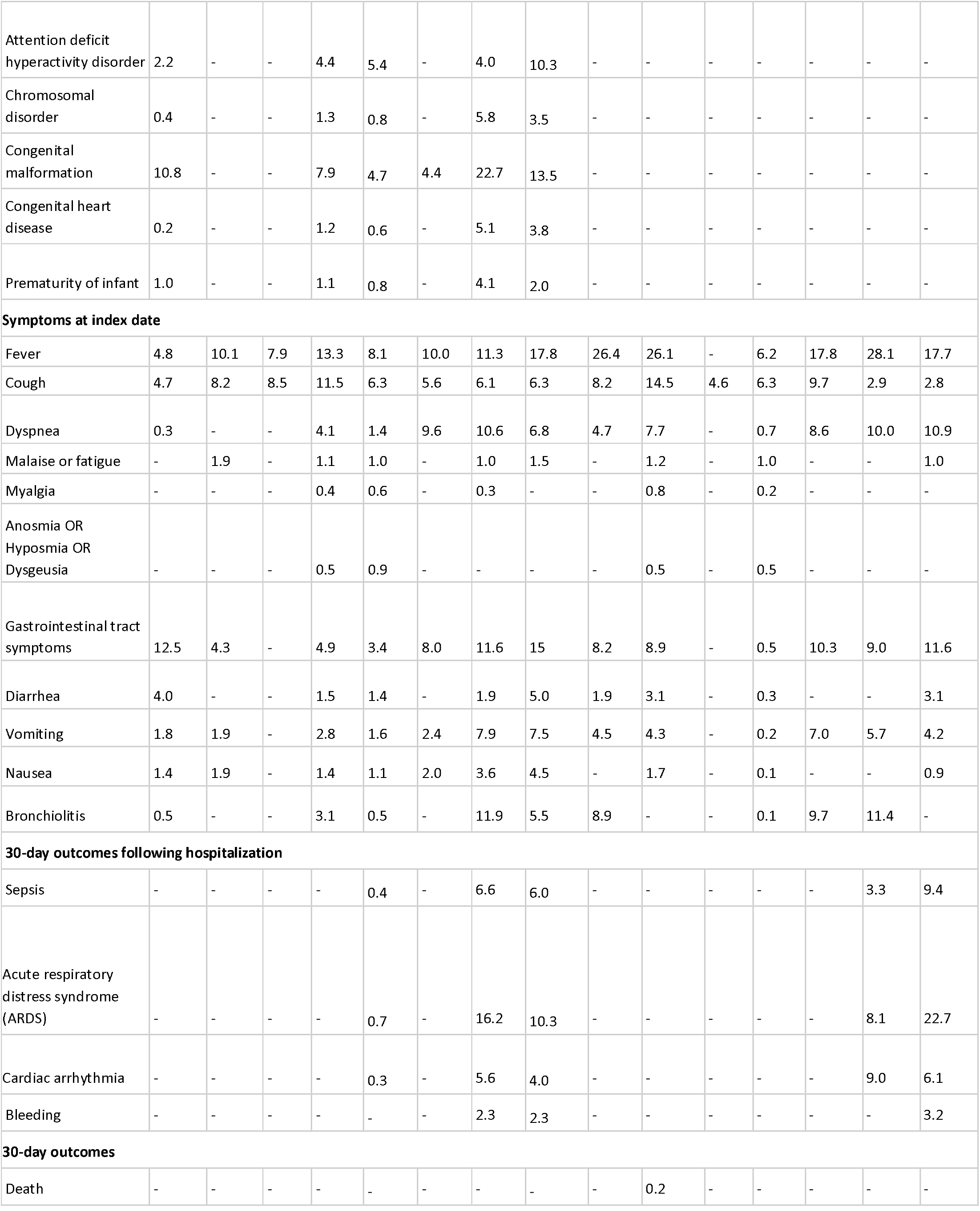
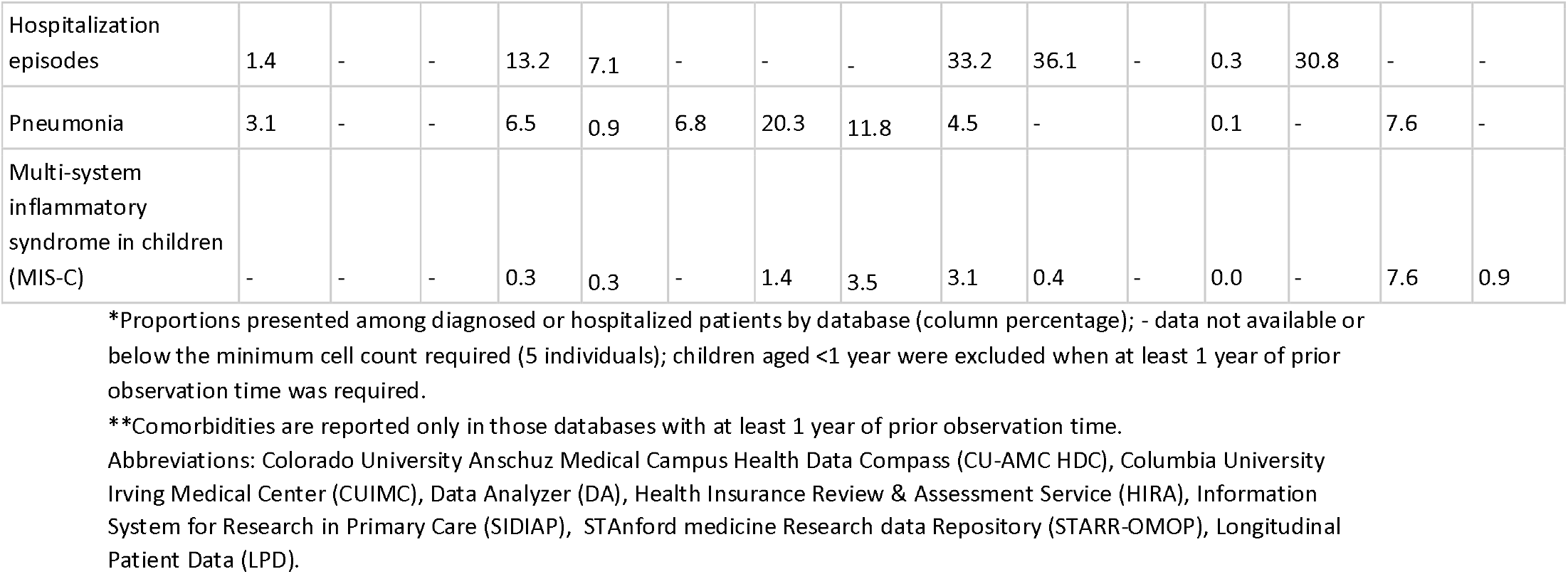
Demographics, comorbidities, symptoms and outcomes among diagnosed and hospitalized COVID-19 children/adolescents (<18 years of age)*

### Comorbidities

Assessed in the year before index date, asthma was the most common baseline comorbidity in patients diagnosed with COVID-19, affecting 10.1% (SIDIAP-Spain) to 33.6% (IQVIA-OpenClaims-US), followed by obesity (from 1.9% in IQVIA-LPD-France to 19.4% in OPTUM-EHR-US). We observed a high prevalence of congenital malformation(s) (7.9% of those diagnosed in IQVIA-OpenClaims-US to 10.8% in SIDIAP-Spain), neurodevelopmental disorders (1.0% of those diagnosed in IQVIA-LPD-France to 8.8% in IQVIA-OpenClaims-US), heart disease (1.2% in IQVIA-LPD-France to 13.1% in IQVIA-OpenClaims-US), type 1 diabetes mellitus (0.2% in SIDIAP-Spain to 0.5% in IQVIA-OpenClaims-US), cancer (0.3% in SIDIAP-Spain to 4% in OPTUM-EHR-US), and chromosomal disorder(s) (0.4% in SIDIAP-Spain to 1.3% in IQVIA-OpenClaims-US). All of these were more common amongst hospitalized children/adolescents with COVID-19 as compared to the diagnosed with COVID-19 cohort (SMD>0.1): 38.8% asthma, 13.1% obesity, 37.0% heart disease (5.1% congenital heart disease), 13.2% cancer, 5.8% chromosomal disorder(s), 22.7% congenital malformation(s), and 4.1% prematurity in IQVIA-OpenClaims-US (Figure 2).

**Figure 2.**
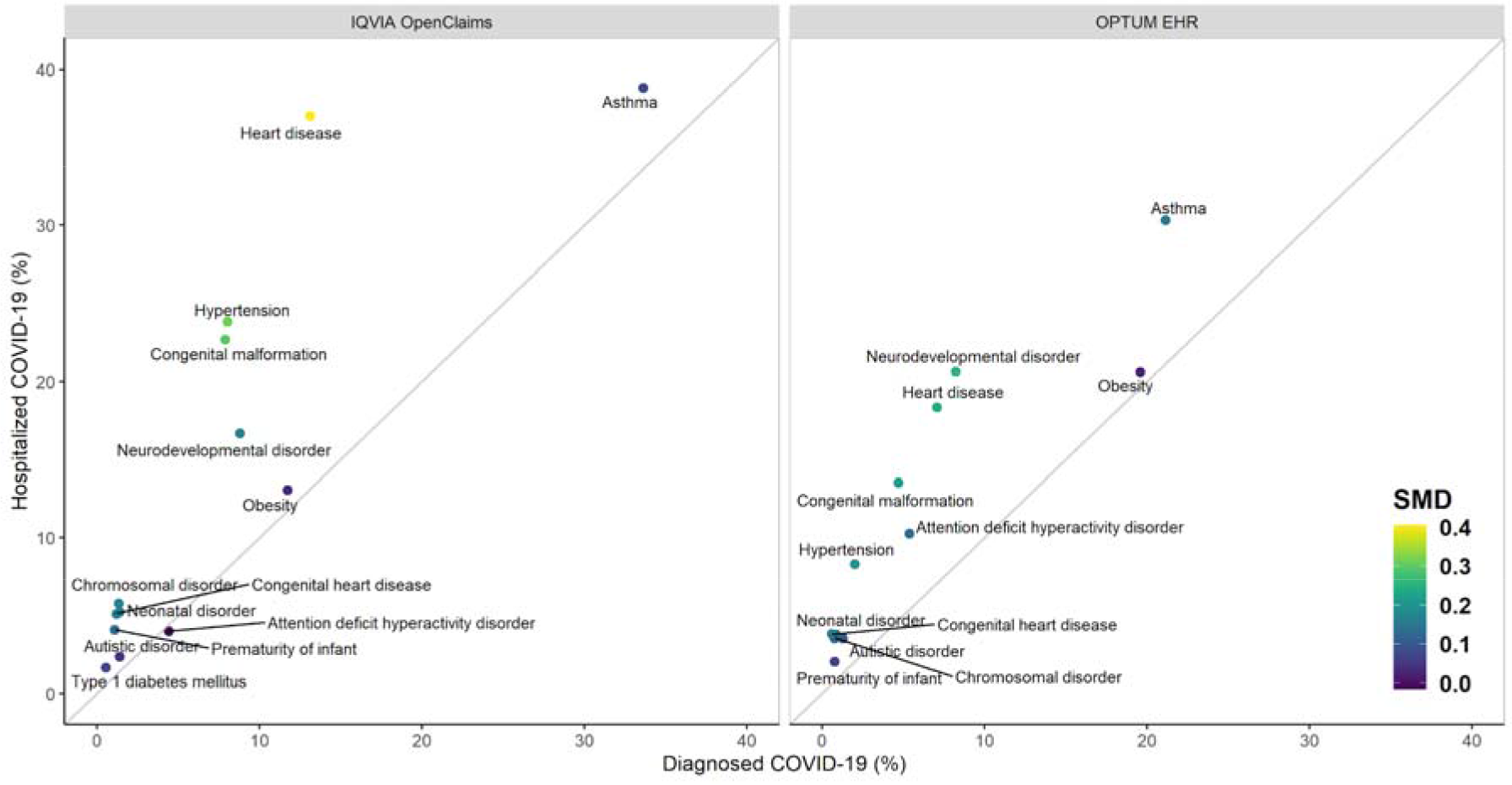
Prevalence of previous comorbidities among children/adolescents (<18 years of age) diagnosed (X axis) compared to hospitalized (Y axis) with COVID-19. SMD = Standardized Mean Differences in prevalence between X and Y.

### Symptoms

Figure 3a shows recorded symptoms at index date for diagnosed vs hospitalized COVID-19 patients while Figure 3b displays symptoms of COVID-19 vs influenza cohorts. The most common reported symptom in COVID-19 was fever, seen in 4.8% (SIDIAP-Spain) to 26.4% (CUIMC-US) of diagnosed cases, and higher (up to 28.1% in CUIMC-US) among hospitalized cases. Second most common was cough, recorded in 4.7% (SIDIAP-Spain) to 14.5% (Premier-US) amongst the diagnosed, and lower (e.g. 2.8% in Premier-US) in hospitalized children/adolescents. Bronchiolitis was recorded in 0.5% (SIDIAP-Spain) to 3.1% (IQVIA-OpenClaims-US) of the diagnosed, and higher (up to 11.9% in Optum EHR-US) in the hospitalized. Gastrointestinal tract symptoms were also common at index date, recorded in 0.5% (HealthVerity-US) to 12.5% (SIDIAP-Spain) in diagnosed, and up to 15.0% (Optum-EHR-US) among hospitalized. Anosmia was <=1% in all participating databases, both in diagnosed and admitted COVID-19 patients. Compared to influenza, COVID-19 diagnosed children/adolescents had less frequently reported symptoms in most databases, with the only exceptions of: dyspnea (e.g. 4.1% in COVID-19 vs 0.4% in influenza in IQVIA-OpenClaims-US), bronchiolitis (e.g. 3.1% in COVID-19 vs 0.2% in influenza in IQVIA-OpenClaims-US), anosmia/hyposmia/dysgeusia (e.g. 0.5% in COVID-19 vs 0.0% in influenza in IQVIA-OpenClaims-US), and gastrointestinal tract symptoms (e.g. 4.9% in COVID-19 vs 3.4% in influenza in IQVIA-OpenClaims-US).

**Figure 3.**
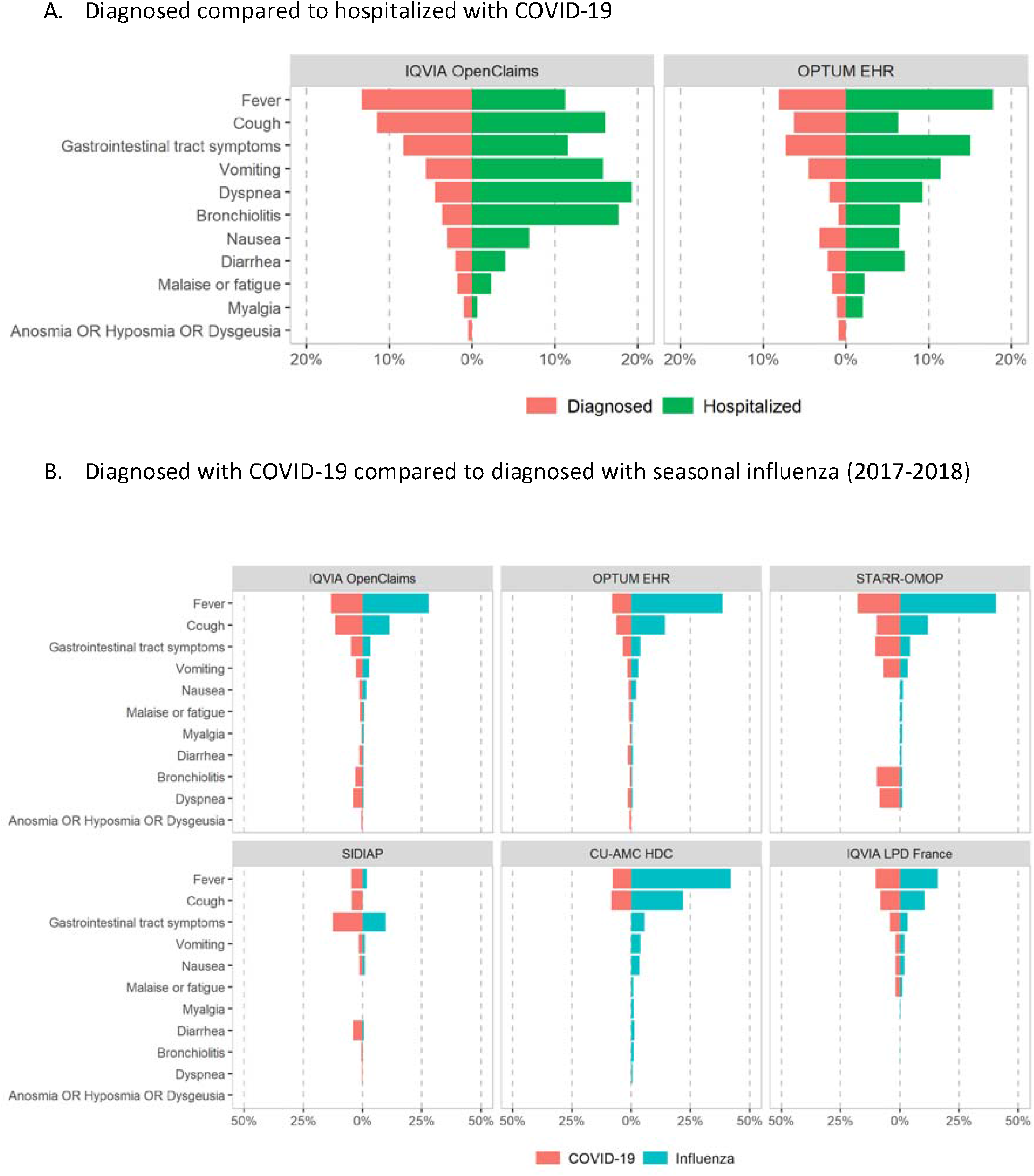
Symptoms recorded at index date among children/adolescents (<18 years of age)

### In-hospital treatments

Use of drugs during hospital admission for COVID-19 amongst children/adolescents is reported in Figures 4a (repurposed) and 4b (adjunctive therapies), based on data from CUIMC-US, OPTUM-EHR-US, Premier-US, and HIRA-South Korea. Repurposed treatments were not commonly used (<10% in all databases), with lopinavir-ritonavir used in <6% in HIRA-South Korea but not in US databases, azithromycin from 4% to 6% in all databases, hydroxychloroquine in 1.5% (Premier-US) to just above 4% (OPTUM-EHR-US), and oseltamivir only in the US, from <1% in Premier-US to just over 5% in CUIMC-US.

**Figure 4.**
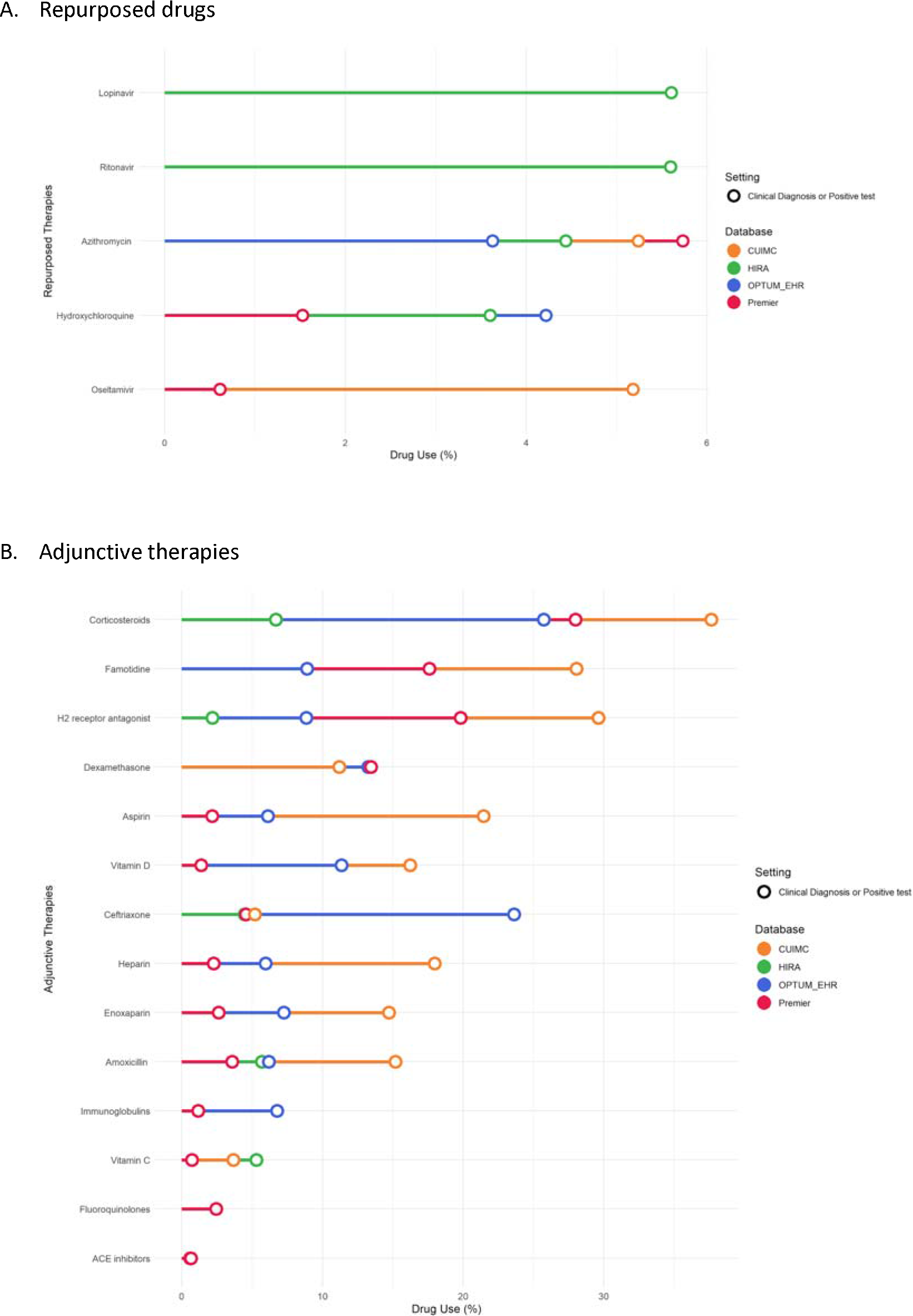
30-day in-hospital use of treatments among children/adolescents (<18 years of age) with COVID-19.

Adjunctive therapies were more common, with systemic corticosteroids used in 6.8% (HIRA-South Korea) to 37.6% (CUIMC-US). Famotidine was the second most common adjunctive treatment, used in 9.0% (OPTUM-EHR-US) to 28.1% (CUIMC-US). Concomitant antithrombotic therapy was also common in the US but not in HIRA-South Korea (no use reported), including aspirin (2.0% in Premier-US to 21.4% in CUIMC-US), heparin (2.2% in Premier-US to 18.1% in CUIMC-US), and enoxaparin (2.8% in Premier-US to 14.8% in CUIMC-US). Antibiotics (ceftriaxone, amoxicillin, fluoroquinolones), vitamin (D and C) supplements, and immunoglobulins were also used with high variability between the contributing databases.

### Health outcomes

Outcomes in the 30-day period following the diagnosis of COVID-19 and hospitalization with COVID-19 are summarized in Table 1 and Figure 5a. Hospitalization was observed in a low proportion (0.3% in HealthVerity-US, 1.4% in SIDIAP-Spain) in the ambulatory/claims databases with testing and testing results data available; in 13.2% in IQVIA-OpenClaims-US; and more frequent (7.1% in OPTUM-EHR-US to 36.1% in Premier-US) in hospital EHR databases. Pneumonia was the most common complication, diagnosed in a wide range from 0.1% (HealthVerity-US) to 6.5% (IQVIA-OpenClaims-US) of those diagnosed, and in 6.8% (HIRA-South Korea) to 20.3% (IQVIA-OpenClaims-US) of those hospitalized with COVID-19. ARDS was the most common in-hospital outcome, affecting 8.1% (CUIMC-US) to 22.7% (Premier-US) of those hospitalized with COVID-19. Sepsis during admission was observed in <2% (N<5; HIRA-South Korea) to 9.4% in Premier-US. Other less common outcomes are reported in Table 1. MIS-C was rare, and seen only in <0.1% (N<5; SIDIAP-Spain) to 0.3% (IQVIA-OpenClaims-US).

**Figure 5.**
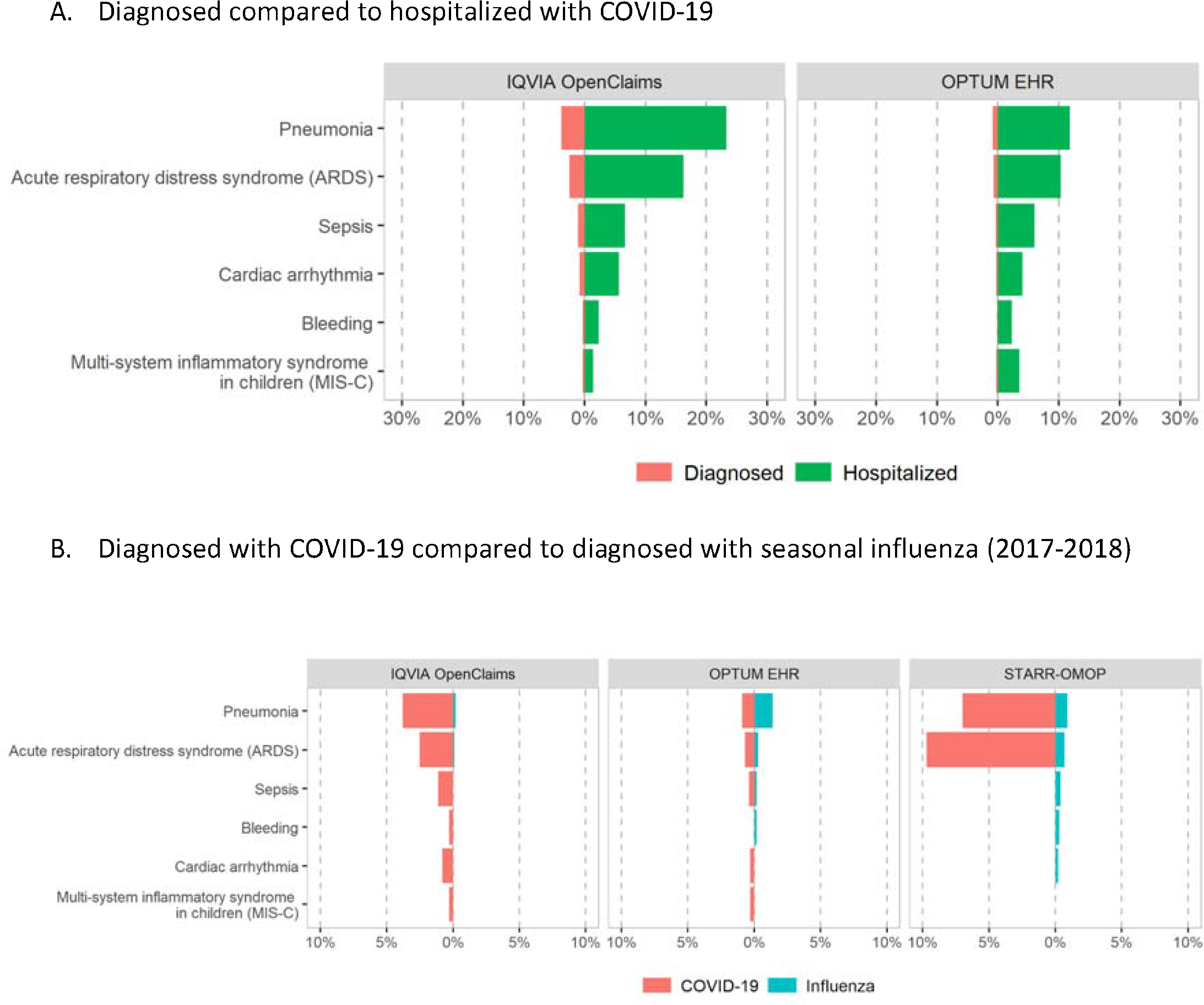
Main 30-day outcomes among children/adolescents (<18 years of age)

A comparison of outcomes in those diagnosed with COVID-19 vs those diagnosed with influenza in previous years is depicted in Figure 5b. Hospitalization rates were higher for COVID-19 vs influenza-diagnosed children/adolescents (e.g. 13.2% vs 0.9% in IQVIA-OpenClaims-US, 33.2% vs 7.4% in CUIMC-US, 30.8% vs 3.7% in STARR-US). Similarly, pneumonia was more common in COVID19-diagnosed participants (e.g. 3.1% vs 0.7% in SIDIAP-Spain, 6.5% vs 2.4% in IQVIA-OpenClaims-US); and so was ARDS (e.g. 2.5% vs 0.1% in IQVIA-OpenClaims-US, 3.3% vs 0.7% in CUIMC-US) and - despite rare-MIS-C (e.g. 0.3% vs 0.0% in IQVIA-OpenClaims-US, 3.1% vs <0.2% in CUIMC-US). Other outcomes had more similar risks in both viral infections (see Figure 5b and Supplementary Table 3).

In a sensitivity analysis, we replicated the analyses including participants who had no prior history available in their EHRs (Supplementary Table 4). Differences in symptoms or outcomes were modest, however it demonstrated the expected incompleteness in prevalent comorbidity.

## DISCUSSION

This study comprehensively reports on the largest cohort of children/adolescents with COVID-19 to date, covering more than 55,000 children/adolescents diagnosed and almost 3,700 hospitalized with COVID-19 in 5 countries from North America, Europe, and Asia. We also studied a cohort of almost two million children/adolescents diagnosed with seasonal influenza in previous years for comparison.

Overall, most cases of COVID-19 diagnosis and related hospitalizations were seen amongst infants and toddlers aged <4 years old, predominantly of male sex. Children/adolescents hospitalized with COVID-19 had a higher prevalence of comorbidities than the overall cohort of those diagnosed with COVID-19, including asthma, obesity, heart disease, cancer, chromosomal disorder(s), and congenital malformation(s). The most commonly observed symptoms were fever and cough, while dyspnea, bronchiolitis, anosmia and gastrointestinal tract symptoms were more common in children/adolescents with COVID-19 than with seasonal influenza, and may aid on the differentiation of COVID-19 from other viral infections.

The drug utilization analysis suggests little use of repurposed drugs and a substantial use of adjunctive therapies among children/adolescents. A high heterogeneity in the use of these drugs across the participating databases underlies differences in COVID-19 management practices across countries among young children/adolescents.

A low proportion of children/adolescents diagnosed with COVID-19 required hospital admission amongst those diagnosed in primary care or outpatient settings (e.g. 1.4% amongst those seen initially in primary care/outpatient), but higher rates (15% to 36%) among those diagnosed in hospital settings.

Hospitalization rates were between 5- and 13-fold higher amongst those children/adolescents diagnosed with COVID-19 vs. those with seasonal influenza in previous years. Fortunately, 30-day fatality following a COVID-19 diagnosis or hospitalization was low: less than 5 children/adolescents died in any of the contributing databases. Respiratory complications (pneumonia and ARDS) were over-represented among children/adolescents diagnosed with COVID-19 compared with seasonal influenza, as was MIS-C. Further outcomes demonstrated similar rates across both cohorts.

### Comparison with other studies

The proportion of children/adolescents under 18 years of age from all observed COVID-19 cases in each database varied from 2.6% in CU-AMC HDC-US to 7.1% in IQVIA-DA-Germany. These values are higher than what has been previously reported in other studies from China (2%) [17], Spain (2%) [18], the US (1.7%) [5], or the UK (0.9%) [19], but in line with the WHO dashboard which reported that children below the age of 14 years old accounted for 3.7% of all COVID-19 cases [1], or the Australian Health Protection Agency which has reported that children/adolescents below the age of 19 years accounted for 4% of confirmed COVID-19 cases in Australia.[20]

Asthma and obesity were the most common baseline comorbidities in children/adolescents with COVID-19; this was expected, and is in keeping with disease prevalence among a general pediatric population.[21] More strikingly, we observed a high prevalence of conditions that are relatively rare in children/adolescents, including congenital malformation/s, neurodevelopmental disorders, heart disease, type 1 diabetes mellitus, cancer, and chromosomal disorder/s. These conditions were more frequent amongst hospitalized children/adolescents with COVID-19 than the overall cohort of those diagnosed with COVID-19. This is in line with previous studies suggesting that children with comorbidity history have higher risk of critical care admission [19,22].

Despite the great variability in the reporting of symptoms across databases, we found that the most commonly identified symptoms in children/adolescents with COVID-19 were consistently fever, cough, dyspnea, and gastrointestinal tract symptoms which are in line with previous studies [7,19,22]. However, the frequency of reported symptoms in our study is generally lower than what has been previously reported in the pediatric literature [7,19,22], suggesting an underestimation in the register of symptoms in the form of structured data in busy actual care settings. An important finding is that COVID-19 diagnosed children/adolescents presented with higher rates of dyspnea, anosmia, and gastrointestinal symptoms than children with seasonal influenza. This information is clinically relevant for differential diagnosis between COVID-19 and influenza among children/adolescents.

We observed great heterogeneity across countries in the use of in-hospital treatments among children/adolescents with COVID-19, which is in line with previous studies in adults [23]. Our analysis suggests little use of antiviral therapies overall, with about 5% of children/adolescents hospitalized with COVID-19 using lopinavir/ritonavir in South Korea (but none in the US); a variable proportion of use of oseltamivir (1% to 5%) in the US (but none in South Korea); a 4-5% of use of azithromycin, and limited use of hydroxychloroquine, ranging from 1.5% to around 4% in both countries. These values are lower than what we previously reported in adults (e.g. use of hydroxychloroquine was 57.9% in the overall population vs. 1.5% in children/adolescents in Premier-US) [23], but in line with recent European cohort studies in hospitalized children/adolescents with COVID-19. A study in the UK reported that 6% (38/591) of hospitalized children/adolescents with COVID-19 received antiviral drugs (30 received acyclovir, 7 received remdesivir, and 3 received chloroquine or hydroxychloroquine) [19], while a study in 25 European countries including 582 children/adolescents found that 7% were treated with hydroxychloroquine, 3% with remdesivir, 1% with lopinavir–ritonavir, 1% with oseltamivir.[7] In contrast, we observed substantial use of different adjunctive therapies; corticosteroids were used in 25-35% in the US but 7% in South Korea. Famotidine (2 to 20%), aspirin (10 to 30%) and vitamin D (2 to 15%) were used in the US but not in South Korea. Antibiotics were also commonly used, with ceftriaxone and amoxicillin amongst the most commonly prescribed. This is consistent with the previous study in in UK where antibiotics were given to 69% (415/601) of hospitalized children with COVID-19.[19]

It is reassuring that occurrence of severe outcomes during the 30 days after diagnosis of COVID-19 was rare in our study, which is in line with previous studies [7,18,19,22]. MIS-C was relatively uncommon, affecting 0.5%-3.1% of all diagnosed cases, but up to 0.9%-7.6% of those hospitalized with COVID-19. These results are in line with previous studies from Europe and the US which have suggested that COVID-19 may be associated with MIS-C in children.[24-26] A separate cohort study found recently that 11% of children with COVID-19 admitted to hospitals in the UK developed MIS-C.[19] These findings are of special relevance given the severity of this condition. Our results also confirmed MIS-C might be specifically related to COVID-19, since it was much less common in patients hospitalized with influenza. Overall, all outcomes were more frequent in children/adolescents with COVID-19 diagnosis than those with a diagnosis of seasonal influenza in 2017-2018, suggesting more severe disease prognosis in children with COVID-19 than influenza. Future research is needed to characterize and determine the long-term outcomes of children/adolescents affected with COVID-19.

### Strengths and limitations

This study has some limitations. First, this study is descriptive in nature. The observed differences between groups should therefore not be interpreted as causal effects. Second, our results are based on secondary data from electronic records collected for administrative and clinical management purposes and re-used for research which may affect the completeness of data recorded and may have erroneous entries, leading to potential misclassification. Such incompleteness could be differential in some instances (e.g. hospital vs primary care settings) and risk information bias for the proposed comparisons. Moreover, variability in coding practices and the heterogeneity in healthcare settings inherent to this study may account for some of the observed variability in the prevalence of comorbidities, symptoms and outcomes. Hospital admissions are also variably recorded in the analyzed databases. The under-reporting of symptoms observed in these data is a key finding of this study, and should be taken into consideration in previous and future similar reports from ‘real-world’ cohorts. Our analysis of mortality is similarly subject to differences by database. For example, data on inpatient deaths are more complete while primary care or outpatient death events are typically imported into a given database from a national or local death register. Finally, the currently available data is restricted to the first six months of 2020 coinciding with the peak of the COVID-19 peak in the studied countries. As data accumulates over time future updates of the results will provide the opportunity to study more recent cohorts of COVID-19 patients, who seem to have a better prognosis overall compared to those diagnosed in the first half of the year.

This study is unique as our approach to characterizing children with an international scope allows for a wide range of variation in healthcare systems and policies against the COVID-19 pandemic. This enables a more complete understanding of the implications of the pandemic for different countries and regions in scope of an international comparison. It also poses a layer of heterogeneity that needs to be considered in the interpretation of our findings, opening a window for new research questions that need to be addressed; particularly around the public health approach for controlling the pandemic spread and severity in children/adolescents and overall. This study represents, to our knowledge, the largest cohort study on pediatric COVID-19 to date, and the only study to provide a comparison with children/adolescents with seasonal influenza in 2017-2018. Our data confirm low rates of complications in children with COVID-19, with severe cases clustering amongst children with previous comorbidities. Despite the large sample size available, MIS-C appears rare.

## Conclusions

COVID-19 affects children/adolescents of all ages but severe outcomes are reassuringly uncommon. There is variability across healthcare systems in different regions of America, Asia and Europe that may explain the observed differences in the epidemiology and clinical management of the disease as well as observed outcomes. Despite negligible fatality, complications including pneumonia, ARDS and MIS-C are more frequent in children/adolescents with COVID-19 than with influenza. Dyspnea, anosmia and gastrointestinal symptoms could help differential diagnosis.

## Data Availability

Analyses were performed locally in compliance with all applicable data privacy laws. Although the underlying data is not readily available to be shared, authors contributing to this paper have direct access to the data sources used in this study. All results (e.g. aggregate statistics, not presented at a patient-level with redactions for minimum cell count) are available for public inquiry. These results are inclusive of site-identifiers by contributing data sources to enable interrogation of each contributing site. All analytic code and result sets are made available at: https://github.com/ohdsi-studies/Covid19CharacterizationCharybdis

## Ethical approval

All the data partners received Institutional Review Board (IRB) approval or exemption. STARR-OMOP had approval from IRB Panel #8 (RB-53248) registered to Leland Stanford Junior University under the Stanford Human Research Protection Program (HRPP). The use of VA data was reviewed by the Department of Veterans Affairs Central IRB, was determined to meet the criteria for exemption under Exemption Category 4(3), and approved for Waiver of HIPAA Authorization. The IRB number for use of HIRA data was AJIB-MED-EXP20-065. The research was approved by the Columbia University Institutional Review Board as an OHDSI network study. The use of SIDIAP was approved by the Clinical Research Ethics Committee of the IDIAPJGol (project code: 20/070-PCV). The use of CPRD was approved by the Independent Scientific Advisory Committee (ISAC) (protocol number 20_059RA2). The use of IQVIA-OpenClaims, IQVIA-LPD-France, and IQVIA-DA-Germany were exempted from IRB approval.

## Transparency declaration

Lead authors affirm that the manuscript is an honest, accurate, and transparent account of the study being reported; that no important aspects of the study have been omitted; and that any discrepancies from the study as planned have been explained.

## Contributorship statement

TDS, DV, KK, APU, PR, DPA conceived and designed the study. SLD, TF, KEL, MEM, KN, JDP, CGR, NHS, PR, KK and TDS coordinated data contributions at their respective sites. AP, AGS, TF, SFB, JDP, KK and TDS analyzed the data; AP produced the figures and tables. TDS, AP, and DPA interpreted the data. TDS, DV, PC, DPA searched the literature and wrote the first draft. WURA, KS, OA, CA, MG, PI, LYHL, TMA, FN and HA reviewed the literature and the manuscript draft. All authors contributed to the revision of the first draft, reviewed and approved the final version of the manuscript.

## Acknowledgements

We would like to acknowledge the patients who suffered from or died of this devastating disease, and their families and carers. We would also like to thank the healthcare professionals involved in the management of COVID-19 during these challenging times, from primary care to intensive care units.

## PPI statement

This research was done without patient involvement. Patients were not invited to comment on the study design and were not consulted to develop patient relevant outcomes or interpret the results. Patients were not invited to contribute to the writing or editing of this document for readability or accuracy.

## Dissemination declaration

We will disseminate the findings of this study to all departments at author-affiliated institutions. In addition, we will further disseminate the findings of this study through press releases, social media, and on institutional web sites upon publication.

---

**Supplementary Table 1.**
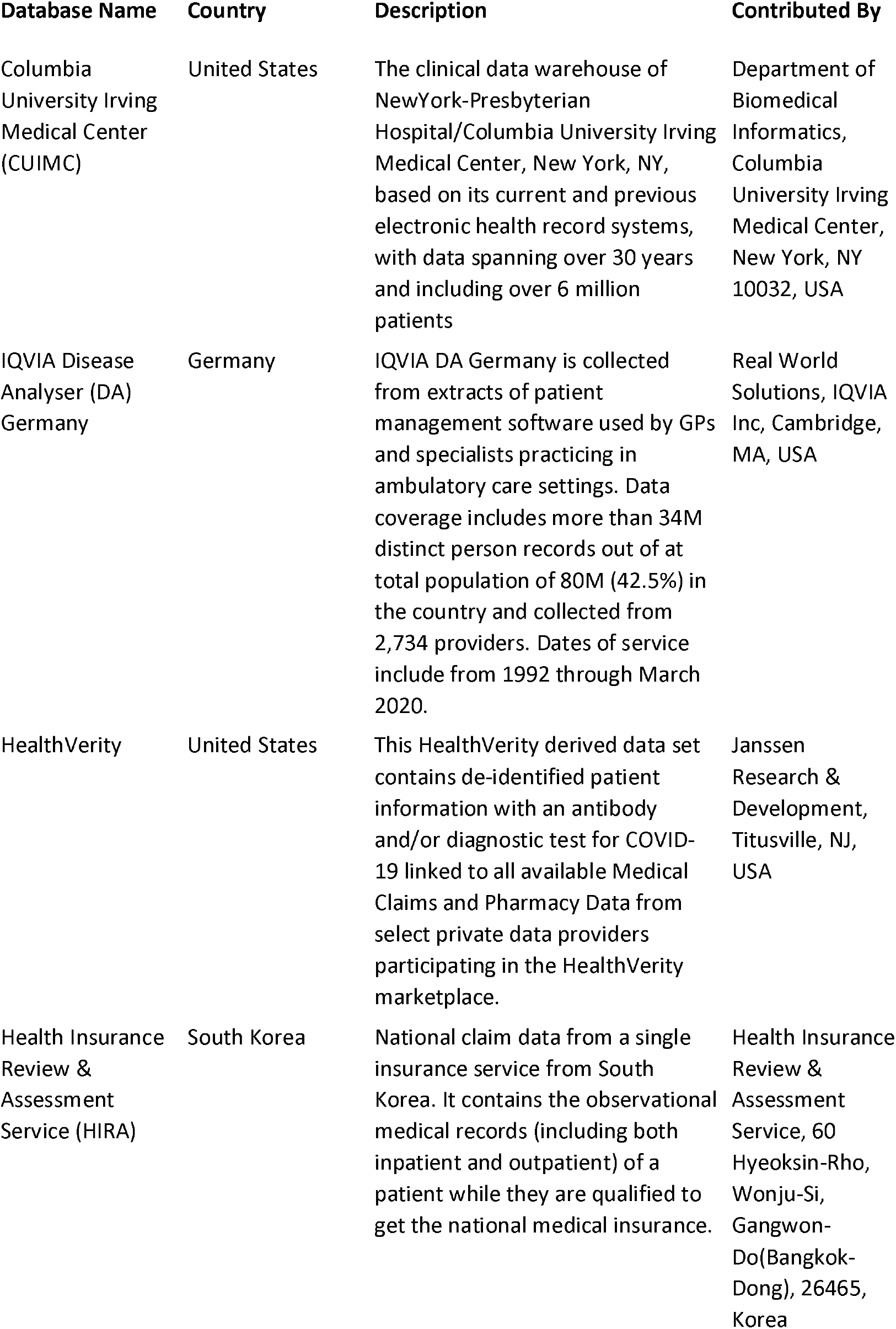
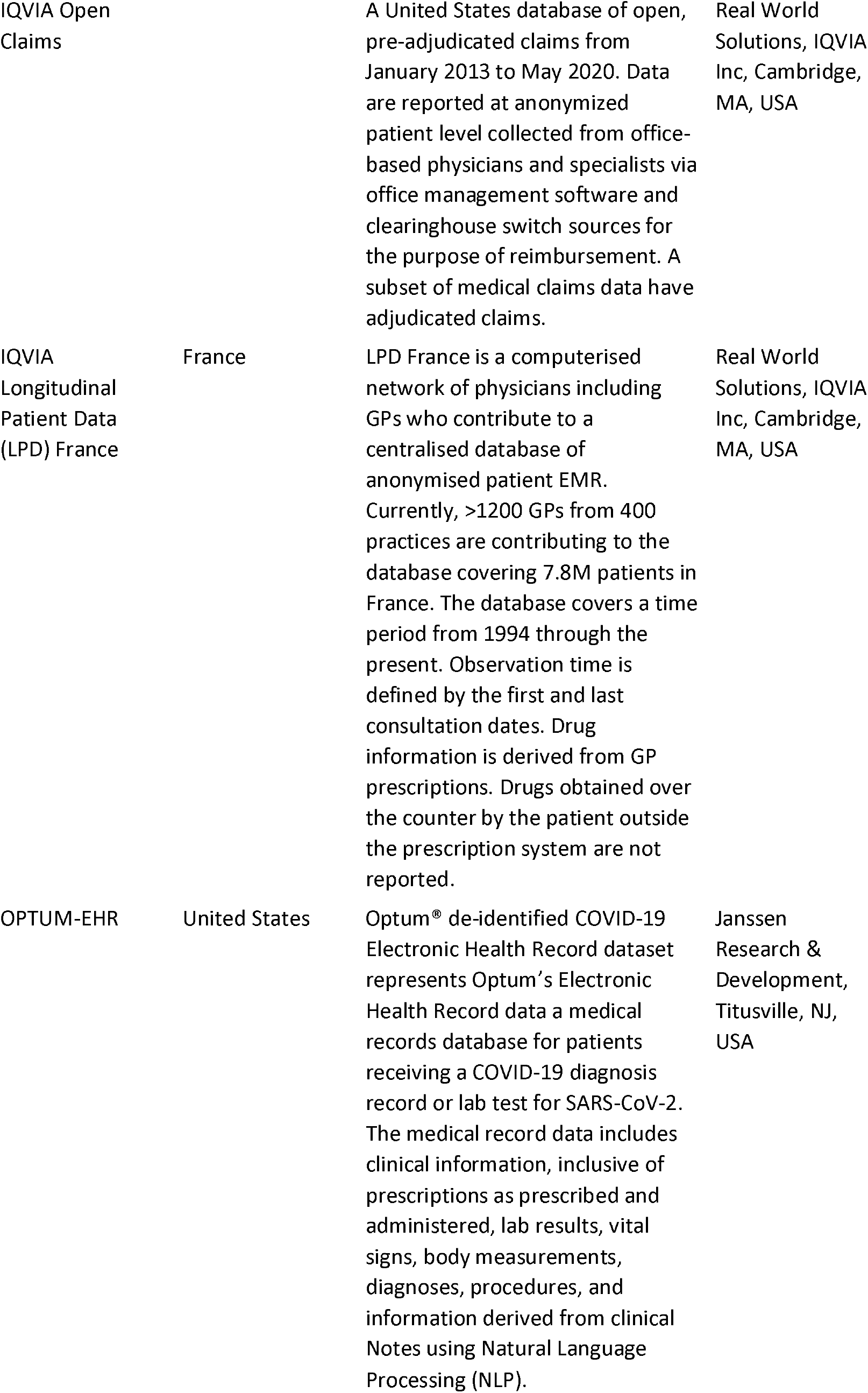
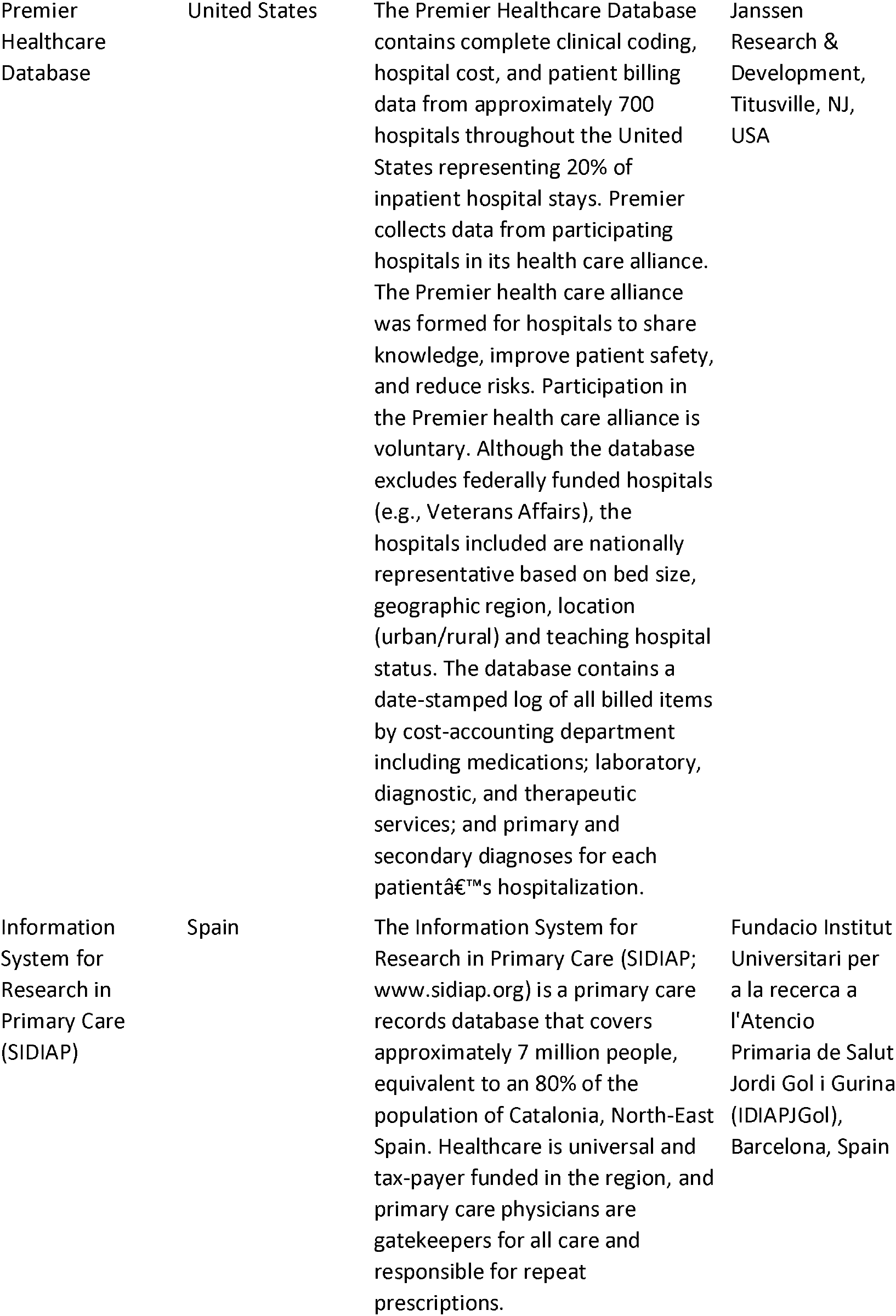
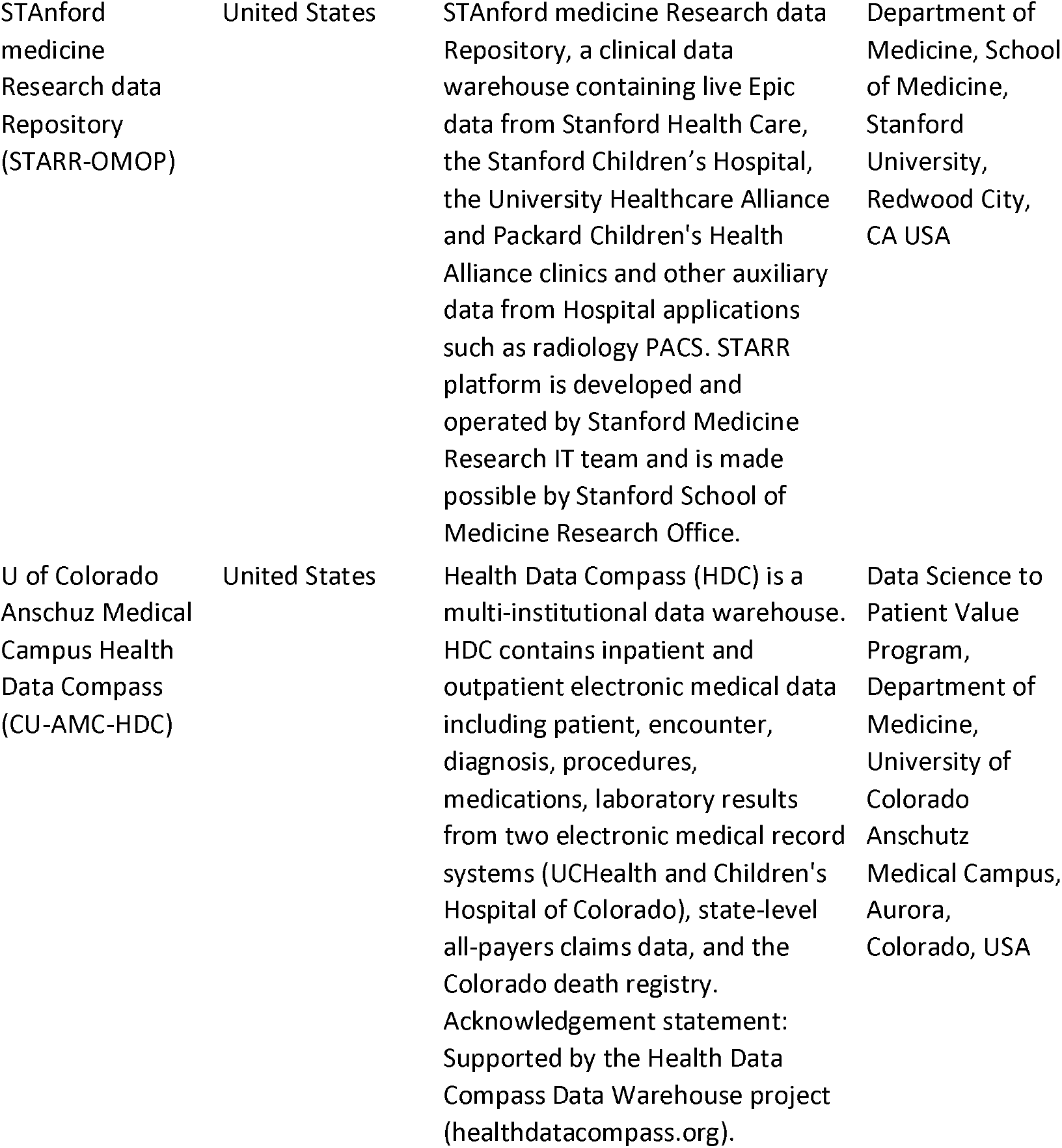
Overview of data sources contributing results.

**Supplementary Table 2.** Cohort definitions and codes.

| Name | Atlas Link |
| --- | --- |
| **<u>COVID-19</u>** |  |
| Persons tested with a COVID-19 diagnosis record or a SARS-CoV-2 positive test with at least 365d prior observation | https://atlas.ohdsi.org/#/cohortdefinition/202 |
| Persons hospitalized with a COVID-19 diagnosis record or a SARS-CoV-2 positive test with at least 365d prior observation | https://atlas.ohdsi.org/#/cohortdefinition/197 |
| Persons tested with a COVID-19 diagnosis record or a SARS-CoV-2 positive test with no required prior observation | http://atlas-covid19.ohdsi.org/#/cohortdefinition/970 |
| Persons hospitalized with a COVID-19 diagnosis record or a SARS-CoV-2 positive test with no required prior observation | http://atlas-covid19.ohdsi.org/#/cohortdefinition/974 |
| **<u>Influenza</u>** |  |
| Persons with Influenza diagnosis or positive test 2017-2018 with at least 365d prior observation | https://atlas.ohdsi.org/#/cohortdefinition/211 |
| Persons hospitalized with influenza diagnosis or positive test 2017-2018 with at least 365d prior observation | https://atlas.ohdsi.org/#/cohortdefinition/212 |
| Persons with Influenza diagnosis or positive test 2017-2018 with no required prior observation | http://atlas-covid19.ohdsi.org/#/cohortdefinition/958 |
| Persons hospitalized with influenza diagnosis or positive test 2017-2018 with no required prior observation | http://atlas-covid19.ohdsi.org/#/cohortdefinition/961 |
| **<u>Comorbidities</u>** |  |
| Asthma | https://atlas.ohdsi.org/#/cohortdefinition/218 |
| Heart disease | https://atlas.ohdsi.org/#/cohortdefinition/231 |
| Hypertension | https://atlas.ohdsi.org/#/cohortdefinition/227 |
| Malignant neoplasm excluding non-melanoma skin cancer | https://atlas.ohdsi.org/#/cohortdefinition/222 |
| Obesity | https://atlas.ohdsi.org/#/cohortdefinition/224 |
| Autistic disorder | https://atlas.ohdsi.org/#/concept/439780 |
| Neonatal disorder | https://atlas.ohdsi.org/#/concept/4042220 |
| Neurodevelopmental disorder | https://atlas.ohdsi.org/#/concept/45771096 |
| Type 1 diabetes mellitus | https://atlas.ohdsi.org/#/concept/201254 |
| Attention deficit hyperactivity disorder | https://atlas.ohdsi.org/#/concept/438409 |
| Chromosomal disorder | https://atlas.ohdsi.org/#/concept/4257441 |
| Congenital malformation | https://atlas.ohdsi.org/#/concept/4079975 |
| Congenital heart disease | https://atlas.ohdsi.org/#/concept/312723 |
| Prematurity of infant | https://atlas.ohdsi.org/#/concept/36675035 |
| **<u>Outcomes</u>** |  |
| **During hospitalization** |  |
| Sepsis during hospitalization | https://atlas.ohdsi.org/#/cohortdefinition/277 |
| Acute Respiratory Distress syndrome (ARDS) during hospitalization | https://atlas.ohdsi.org/#/cohortdefinition/278 |
| Cardiac arrhythmia | https://atlas.ohdsi.org/#/cohortdefinition/248 |
| Bleeding | https://atlas.ohdsi.org/#/cohortdefinition/238 |
| **30-day outcomes** |  |
| Death | http://atlas-covid19.ohdsi.org/#/cohortdefinition/166 |
| Hospitalization episodes | http://atlas-covid19.ohdsi.org/#/cohortdefinition/917 |
| Pneumonia | http://atlas-covid19.ohdsi.org/#/cohortdefinition/938 |
| Multi-system inflammatory syndrome in children (MIS-C) | http://atlas-covid19.ohdsi.org/#/cohortdefinition/940 |

**Supplementary Table 3.** Demographics, comorbidities, symptoms and outcomes among diagnosed with seasonal influenza (2017-2018) among children/adolescents aged below 18 years*.

|  | At least 1 year of prior observation |  |  |  |  | No prior observation time |  |  |  |
| --- | --- | --- | --- | --- | --- | --- | --- | --- | --- |
|  | SIDIAP<br>(Spain) | IQVIA<br>LPD<br>(France) | CU-AMC<br>HDC<br>(US) | IQVIA<br>OpenClai<br>ms (US) | OPTUM<br>EHR (US) | CUIMC<br>(US) | IQVIA DA<br>(German<br>y) | HealthVe<br>rity (US) | STARR-OMOP<br>(US) |
|  | n=26929 | n=22425 | n=3339 | n=187348<br>0 | n=3665 | n=2112 | n=19863 | n=502 | n=2831 |
| **Age (years)** |  |  |  |  |  |  |  |  |  |
| 00-04 | 22.0 | 20.8 | 23.7 | 24.4 | 24.1 | 40.2 | 29.3 | 21.7 | 25.3 |
| 05-09 | 36.7 | 35.3 | 35.6 | 37.5 | 31.3 | 36.2 | 35.7 | 29.7 | 34.7 |
| 10-14 | 29.2 | 27.0 | 27.0 | 26.3 | 24.9 | 17.2 | 23.3 | 23.7 | 27.8 |
| 15-19 | 12.0 | 17.0 | 13.6 | 11.8 | 19.7 | 6.5 | 11.7 | 24.9 | 12.2 |
| **Gender** |  |  |  |  |  |  |  |  |  |
| Female | 47.5 | 47.8 | 49.6 | 48.5 | 50.9 | 48.2 | 47.4 | 50.6 | 46.2 |
| Male | 52.5 | 51.9 | 50.4 | 51.5 | 49.1 | 51.8 | 52.6 | 49.4 | 53.8 |
| **Comorbidities\*\*** |  |  |  |  |  |  |  |  |  |
| Autistic disorder | 0.5 | - | 0.9 | 1.0 | 1.4 | - | - | - | - |
| Neonatal disorder | 2.4 | - | 0.9 | 0.3 | 0.9 | - | - | - | - |
| Neurodevelopmental disorder | 6.7 | 0.6 | 3.5 | 6.4 | 10.2 | - | - | - | - |
| Asthma | 8.6 | 16.4 | 15.9 | 27.5 | 29.7 | - | - | - | - |
| Obesity | 8.3 | 1.5 | 3.4 | 4.2 | 13.9 | - | - | - | - |
| Heart disease | 1.8 | 0.4 | 2.3 | 4.5 | 7.2 | - | - | - | - |
| Malignant neoplasm excluding non-melanoma skin cancer | 0.1 | 0.1 | - | 0.5 | 3.5 | - | - | - | - |
| Hypertension | 0.1 | 0.1 | 0.4 | 1.1 | 1.2 | - | - | - | - |
| Type 1 diabetes mellitus | 0.1 | 0.0 | - | 0.2 | 0.4 | - | - | - | - |
| Attention deficit hyperactivity disorder | 2.4 | 0.0 | 1.7 | 4.5 | 6.7 | - | - | - | - |
| Chromosomal disorder | 0.1 | - | 0.4 | 0.3 | 0.8 | - | - | - | - |
| Congenital malformation | 11.2 | 0.4 | 2.5 | 2.7 | 4.7 | - | - | - | - |
| Congenital heart disease | 0.2 | - | 0.3 | 0.3 | 0.6 | - | - | - | - |
| Prematurity of infant | 0.6 | - | 0.4 | 0.2 | 0.6 | - | - | - | - |
| **Symptoms at index date** |  |  |  |  |  |  |  |  |  |
| Fever | 1.7 | 15.9 | 42.1 | 27.9 | 38.6 | 51.8 | 9.3 | 31.3 | 40.6 |
| Cough | 0.2 | 10.4 | 21.9 | 11.4 | 14.4 | 16 | 4.1 | 16.9 | 11.8 |
| Dyspnea | 0.0 | 0.0 | 0.6 | 0.4 | 0.6 | 2.2 | 0.9 | - | 1.1 |
| Malaise or fatigue | 0.0 | 1.0 | 0.9 | 0.7 | 0.7 | 0.5 | 0.4 | - | 1.1 |
| Myalgia | 0.0 | 0.3 | 1.0 | 0.6 | 0.5 | 0.5 | 0.1 | - | 1.0 |
| Anosmia OR<br>Hyposmia OR<br>Dysgeusia | - | - | - | 0.0 | - | - | - | - | - |
| Gastrointestinal tract symptoms | 9.6 | 3.2 | 5.6 | 3.4 | 3.9 | 6.1 | 2.3 | 5.4 | 4.5 |
| Diarrhea | 0.6 | 0.0 | 1.3 | 0.5 | 0.8 | 0.9 | - | - | 0.8 |
| Vomiting | 1.1 | 2.0 | 3.9 | 2.8 | 2.9 | 4.9 | 1.9 | 4.4 | 3.4 |
| Nausea | 1.1 | 2.0 | 3.5 | 1.6 | 2.1 | 0.6 | 2.0 | 3.2 | 1.4 |
| Bronchiolitis | 0.2 | 0.2 | 1.0 | 0.5 | 0.4 | 0.8 | 0.0 | - | 1.1 |
| **30-day outcomes during hospitalization** |  |  |  |  |  |  |  |  |  |
| Sepsis | - | - | - | - | 0.2 | 0.4 | - | - | 0.4 |
| Acute respiratory distress syndrome (ARDS) | - | - | - | 0.1 | 0.3 | 0.7 | - | - | 0.7 |
| Cardiac arrhythmia | - | - | - | - | - | 0.7 | - | - | 0.2 |
| Bleeding | - | - | - | - | 0.2 | 0.6 | - | - | 0.3 |
| **30-day outcomes** |  |  |  |  |  |  |  |  |  |
| Death | - | - | - | - | - | - | - | - | - |
| Hospitalization episodes | - | - | 1.6 | 0.9 | 18.7 | 7.4 | - | 1.4 | 3.7 |
| Pneumonia | 0.7 | 0.4 | 2.2 | 2.4 | 1.4 | 3.1 | 2.0 | - | 2.4 |
| Multi-system inflammatory syndrome in children (MIS-C) | - | 0.0 | - | 0.0 | - | - | 0.0 | - | - |

\*Proportions presented among diagnosed or hospitalized patients by database (column percentage); - data not available or below the minimum cell count required (5 individuals); children aged <1 year were excluded when at least 1 year of prior observation time was required.
\*\*Comorbidities are reported only in those databases with at least 1 year of prior observation time.
Abbreviations: Colorado University Anschutz Medical Campus Health Data Compass (CU-AMC HDC), Columbia University Irving Medical Center (CUIMC), Data Analyzer (DA), Health Insurance Review & Assessment Service (HIRA), Information System for Research in Primary Care (SIDIAP), STANford medicine Research data Repository (STARR-OMOP), Longitudinal Patient Data (LPD).

**Supplementary Table 4.** Characteristics of diagnosed and hospitalized COVID-19 children/adolescents with no prior observation time in those databases with available observation time*.

|  | Diagnosed |  |  |  |  | Hospitalized |  |  |
| --- | --- | --- | --- | --- | --- | --- | --- | --- |
|  | SIDIAP<br>(Spain) | IQVIA<br>LPD<br>(France) | CU-AMC<br>HDC<br>(US) | IQVIA<br>OpenClaims<br>(US) | OPTUM<br>EHR<br>(US) | HIRA<br>(South<br>Korea) | IQVIA<br>OpenClaims<br>(US) | OPTUM<br>EHR<br>(US) |
|  | n =<br>5037 | n = 979 | n =<br>286 | n = 17618 | n =<br>7737 | n = 251 | n = 3055 | n = 748 |
| **Comorbidities** |  |  |  |  |  |  |  |  |
| Autistic disorder | 0.7 | - | - | 0.4 | 0.3 | - | 0.9 | 1.9 |
| Neonatal disorder | 1.7 | - | - | 1.9 | 0.8 | - | 7.8 | 5.7 |
| Neurodevelopmental disorder | 6.1 | - | - | 1.4 | 1.6 | - | 3.4 | 7.8 |
| Asthma | - | - | - | 0.9 | 0.7 | - | 1.4 | 2.8 |
| Obesity | - | - | - | 0.1 | 0.5 | - | 0.4 | 1.6 |
| Heart disease | - | - | - | 0.6 | 0.5 | - | 1.7 | 3.6 |
| Malignant neoplasm excluding non-melanoma skin cancer | - | - | - | 0.1 | 0.1 | - | 0.2 | 1.2 |
| Hypertension | - | - | - | 0.1 | 0.1 | - | 0.5 | 0.9 |
| Type 1 diabetes mellitus | 0.2 | - | - | 0.3 | 0.2 | - | 0.8 | 0.8 |
| Attention deficit hyperactivity disorder | 1.8 | - | - | 0.6 | 1.0 | - | 1.0 | 3.7 |
| Chromosomal disorder | 0.3 | - | - | 0.7 | 0.3 | - | 2.9 | 2.1 |
| Congenital malformation | 9.2 | - | - | 2.6 | 1.7 | - | 10.4 | 8.7 |
| Congenital heart disease | 0.2 | - | - | 0.6 | 0.3 | - | 2.8 | 1.9 |
| Prematurity of infant | 0.8 | - | - | 0.5 | 0.5 | - | 2.6 | 2.9 |
| **Symptoms at index date** |  |  |  |  |  |  |  |  |
| Fever | 5.2 | 9.6 | 8.7 | 14.7 | 9.9 | 10 | 12.2 | 23.7 |
| Cough | 4.5 | 7.5 | 9.1 | 11.5 | 6.6 | 5.6 | 6.2 | 8.0 |
| Dyspnea | 0.3 | - | 4.5 | 4.6 | 1.7 | 9.6 | 11.2 | 7.0 |
| Malaise or fatigue | - | 1.6 | - | 1.1 | 1.0 | - | 1.0 | 1.3 |
| Myalgia | - | - | - | 0.3 | 0.6 | - | 0.2 | 1.2 |
| Anosmia OR Hyposmia OR Dysgeusia | - | - | - | 0.4 | 0.7 | - | - | <0.7 |
| Gastrointestinal tract symptoms | 11.8 | 3.7 | - | 5.4 | 3.9 | 8.0 | 10.9 | 14.0 |
| Diarrhea | 3.7 | - | - | 1.6 | 1.7 | - | 2.4 | 5.1 |
| Vomiting | 1.7 | 1.6 | - | 3.2 | 1.9 | 2.4 | 7.5 | 7.8 |
| Nausea | 1.4 | 1.6 | - | 1.2 | 1.1 | 2.0 | 2.6 | 3.5 |
| Bronchiolitis | 1.3 | - | - | 6.6 | 1.1 | - | 21.9 | 8.0 |
| **30-day outcomes** |  |  |  |  |  |  |  |  |
| Death | - | - | - | - | - | - | - | - |
| Hospitalization episodes | 0.5 | - | 3.8 | 12.6 | 6.4 | 100 | 97.7 | 100 |
| Pneumonia | 0.1 | - | - | 4.0 | 1.0 | 6.4 | 19.5 | 10.2 |
| Multi-system inflammatory syndrome in children (MIS-C) | - | - | - | 0.2 | 0.5 | - | 1.0 | 3.7 |
| **30-day outcomes during hospitalization** |  |  |  |  |  |  |  |  |
| Sepsis | - | - | - | 1.2 | 0.7 | - | 6.0 | 7.9 |
| Acute respiratory distress syndrome (ARDS) | - | - | - | 3.1 | 1.0 | - | 16.1 | 10.7 |
| Cardiac arrhythmia | - | - | - | 0.9 | 0.3 | - | 5.0 | 2.9 |
| Bleeding | - | - | - | 0.3 | 0.3 | - | 1.9 | 3.3 |

\*Proportions presented among diagnosed or hospitalized patients by database (column percentage); - data not available or below the minimum cell count required (5 individuals).
\*\*Comorbidities are reported only in those databases with at least 1 year of prior observation time.
Abbreviations: Colorado University Anschutz Medical Campus Health Data Compass (CU-AMC HDC), Columbia University Irving Medical Center (CUIMC), Data Analyzer (DA), Health Insurance Review & Assessment Service (HIRA), Information System for Research in Primary Care (SIDIAP), STANford medicine Research data Repository (STARR-OMOP), Longitudinal Patient Data (LPD).

